# Effect of cardiorespiratory exercise during rehabilitation on functional recovery early post-stroke: a cohort study

**DOI:** 10.1101/2024.08.09.24311772

**Authors:** Sarah Thompson, Augustine J. Devasahayam, Cynthia J. Danells, David Jagroop, Elizabeth L. Inness, Avril Mansfield

**Author notes:** Corresponding author: Avril Mansfield; 550 University Ave, Toronto, ON, Canada, M5G 2A2.

## Abstract

**Background:** People with stroke often have low cardiorespiratory fitness, hindering daily activities and rehabilitation participation. Cardiorespiratory exercise (CRE) early post-stroke can improve fitness, facilitating participation in rehabilitation, and may promote neuroplasticity. This longitudinal observational study aimed to determine the effect of CRE during routine inpatient stroke rehabilitation on motor and cognitive function, functional ambulation, and motor impairment.

**Methods:** Data were collected from charts of patients (n=504) admitted to two rehabilitation hospitals in Ontario, Canada, over 14 month periods. Patients were classified into three groups: ‘Prescribed’, ‘Incidental’, or no cardiorespiratory exercise (‘None’). Functional independence Measure (FIM), Functional Ambulation Category (FAC), and Chedoke-McMaster Stroke Assessment (CMSA) scores were compared between groups at discharge from rehabilitation, controlling for age, length of stay, and scores at baseline.

**Results:** Patients who had cardiorespiratory exercise included in their treatment plan (i.e., Prescribed group) had higher FIM total and motor sub-scores at discharge than the None group (Site A; FIM total mean between-group difference: 13.2, p<0.0001; FIM motor mean between-group difference: 13.1, p<0.0001), or than those that completed cardiorespiratory exercise without a prescription (‘Incidental’ group; Site B; FIM Total mean between-group difference: 13.6, p=0.031; FIM motor mean between-group difference: 12.9, p=0.010). At both sites, FIM cognitive sub-scores and CMSA leg scores were higher at discharge for the Prescribed group than the None group (FIM cognitive mean between-group difference: 1.2, p=0.038; CMSA leg mean between-group difference: 0.5, p=0.0099). FAC scores were higher at discharge for the two exercise groups compared to the group that did not complete cardiorespiratory exercise at Site A only (p=0.0010).

**Conclusions:** Findings support that cardiorespiratory exercise as part of routine in-patient rehabilitation early post-stroke is associated with improved functional independence and ambulation. However, the observational design limits causal inferences, highlighting the need for controlled studies to confirm cardiorespiratory exercise benefits in early stroke recovery.

## INTRODUCTION

People with stroke typically have low cardiorespiratory fitness, which can hinder participation in stroke rehabilitation and lead to difficulty in completing daily activities.^1^ Cardiorespiratory exercise (CRE) is planned, structured, repetitive physical activity performed at a prescribed frequency, intensity, and duration with the goal of improving or maintaining cardiorespiratory fitness.^2^ CRE can improve cardiorespiratory fitness early post-stroke,^3^ and in turn, may facilitate functional recovery and participation in daily and rehabilitation activities.^1,4^ CRE is also thought to promote neuroplasticity, through increased levels of signaling molecules like brain-derived neurotrophic factor (BDNF) and insulin-like growth factor-1 (IGF-1), and increased synaptogenesis.^5,6^ The greatest amount of recovery takes place in the sub-acute phase of stroke recovery (≤6 months),^1,7^ so this phase is a critical time for CRE participation to enhance plasticity. Indeed, evidence suggests that CRE early post-stroke is safe and effective for improving cardiorespiratory fitness.^3,4,8^ Because of the benefits of cardiorespiratory exercise post-stroke, it is included in the Canadian stroke best practice guidelines.^9^

Despite the benefits of CRE early post-stroke, physiotherapists may prioritize functional activities during the limited time they have with their patients during stroke rehabilitation, rather than working on activities, like CRE, that target physiological impairments underlying function.^10^ Previous research demonstrated that people with stroke who complete cardiorespiratory exercise improved mobility^11^ and exhibited global cognition improvements.^12^ Most studies in these reviews included people with chronic stroke,^11,12^ whereas the greatest gains in physical and cognitive function occur in the sub-acute phase.^13,14^ Improvements in physical function and evidence of cognitive gains due to CRE suggests that recovery outcomes such as functional independence, mobility, and motor impairment are expected to improve in those who complete cardiorespiratory exercise in the first few months post-stroke.

The purpose of this longitudinal observational study is to determine if participation in CRE early post-stroke during routine inpatient stroke rehabilitation is associated with improved recovery of (1) motor and cognitive function (as measured using the Functional Independence Measure; FIM^TM^),^15^ (2) independent ambulation (as measured using the Functional Ambulation Categories; FAC),^16^ and (3) motor impairment (as measured using the Chedoke McMaster Stroke Assessment; CMSA).^17^ Our primary outcome is the FIM as this is a key determinant of stroke rehabilitation service provision and discharge planning within many healthcare systems, including Ontario.^18,19^ Safe independent ambulation, as assessed using the FAC, was chosen as a secondary outcome as patients post-stroke typically prioritize mobility in their goals for rehabilitation.^20^ Global effects of cardiorespiratory exercise early post-stroke, such as improvements in motor impairments,^21^ may be captured through changes in CMSA motor scores from the affected upper and lower extremities. Our primary hypothesis is that the total FIM scores will improve more from admission to discharge from in-patient stroke rehabilitation for those who completed CRE than those who did not. Our secondary hypotheses are that FIM motor and cognitive sub-scores, FAC, and CMSA leg, foot, arm, and hand motor scores will improve more for those who completed cardiorespiratory exercise during stroke rehabilitation than those who did not. We employed an observational design to determine the effect of any CRE completed as part of routine stroke rehabilitation on functional ambulation and impairment outcomes at two rehabilitation hospitals. This study design allowed us to observe the effects of CRE implemented during routine practice, rather than within the context of a clinical trial.

## METHODS

### Design

This longitudinal observational study is a secondary analysis of data collected from two of four sites participating sites in a convergent mixed methods study in Ontario, Canada.^10^ The study was approved by the research ethics boards of both sites (University Health Network protocol number: 20-5695, Sunnybrook Research Institute protocol number: 3605), and is reported according to the STROBE guidelines.^22^

### Participants

We conducted retrospective chart reviews of all patients with diagnosed stroke admitted to inpatient rehabilitation at two rehabilitation hospitals (Sunnybrook St. John’s Rehabilitation Hospital - Site A and Toronto Rehabilitation Institute, University Centre – Site B); all patients admitted to the hospitals during the pre-determined time-periods were included in the study. Chart reviews covered 14-months, 2 months prior to and 12 months after the start of the coronavirus disease 2019 (COVID-19) pandemic. The months of March and October of 2019 were selected to avoid disruptions in care due to staff vacation and statutory holidays in the summer and winter months. Chart reviews during the COVID-19 pandemic were conducted from June 2022 to May 2023 for Site A and from January 2021 to January 2022 for Site B. Patients were excluded from the analysis if their length of stay was ≤7 days and/or if FIM scores or FAC were missing at admission and/or discharge. A waiver of patient consent for chart review was approved,^23^ although patients in the post-pandemic cohorts were informed of the study and had the opportunity to opt out of chart review. Including all patients admitted during the review period helped to remove bias due to selection of only those patients who are willing and able to consent to participate.

### Outcomes

The following variables were extracted from patients’ electronic or physical medical charts: cohort descriptors including age, sex; stroke information (time post-stroke, affected side, lesion location, type of stroke); functional outcomes (FIM, FAC); motor recovery (CMSA arm, hand, leg, and foot scores); and details of any CRE completed during rehabilitation.

Patients were stratified into cohorts (pre- and post-pandemic) and classified into three groups: (1) patients for whom CRE was included in their treatment plan and they completed CRE at least once during rehabilitation (‘Prescribed’ group); (2) patients who completed ‘incidental’ CRE, meaning they used a piece of CRE equipment (e.g., recumbent stepper, treadmill) during physiotherapy sessions, but CRE was not included in their treatment plan (‘Incidental’ group); and (3) patients who completed no CRE (‘None’ group). For the Prescribed and Incidental groups, average CRE session duration was calculated as the mid-point between the minimum and maximum session duration. Time spent exercising per day was calculated as the average CRE session duration multiplied by the number of sessions and divided by length of stay.

### Data analysis

Statistical analyses were performed using SAS (Version 9.4, SAS Institute, Inc., Cary, North Carolina, USA). Patients with missing data for a specific variable were excluded from analysis of that variable. Three-way analysis of variance (ANOVA) was used to analyse continuous cohort descriptors and outcomes at baseline, with factors of site, cohort (pre-versus post-pandemic), and group, as well as the corresponding interaction effects. Chi-square test was used to compare categorical cohort descriptors between sites and groups. We conducted three-way analysis of covariance (ANCOVA) to determine the effect of participation in cardiorespiratory exercise during stroke rehabilitation on primary and secondary outcomes. Site, cohort, and group, and the corresponding interaction effects were factors in the ANCOVA; age, length of stay, and the dependent variable value at admission to rehabilitation were included as covariates. The following dependent variables were analysed: total FIM scores, FIM motor and cognitive sub-scores, FAC, and CMSA leg, foot, arm, and hand motor impairment scores. In the ANOVA and ANCOVAs, there were no significant site-by-cohort-by-group interaction effects, nor were there any significant site-by-cohort or cohort-by-group interaction effects. Therefore, to simplify the analysis, we repeated the analyses as described above using two-way ANOVAs and ANCOVAs, with the cohort factor and associated interaction effects removed. When there were significant site-by-group interaction effects (p<0.05) Tukey’s post-hoc testing was used to determine the statistically significant effects (α=0.05).

For the two exercise groups (Prescribed and Incidental), we conducted multiple linear regression to determine the relationship between total time spent exercising and change in outcomes (FIM total score, FIM motor and cognitive scores, FAC, and CMSA scores). Change scores were calculated as the discharge score minus the admission score, divided by length of stay. Site, age, and time spent exercising were dependent variables in the multiple regression.

## RESULTS

There were 504 patients included in the analysis across both sites and cohorts; Table 1 shows patient characteristics at admission to in-patient stroke rehabilitation. There were differences between sites in CRE practices, and changes in CRE practices from pre- to post-pandemic cohorts. Pre-pandemic, both sites had a similar proportion of patients in the Prescribed group (Site A: 14/45; Site B 13/41); however, Site A had more patients in the Incidental group than in Site B (Site A: 26/45; Site B: 4/41). Post-pandemic, more patients participated in CRE at Site A than Site B.

**Table 1:** Patient characteristics on admission to rehabilitation. Values are mean (standard deviation) for continuous variables, or count (% of group) for categorical variables. The p-values are for the analysis of covariance (continuous variables) or chi-square test (categorical variables). When there were significant group-by-site interaction effects, significant differences from post-hoc testing are indicated with superscript letters; group-site combinations with the same letter were statistically significantly different from each other.

|  | Site A |  |  |  |  |  | Site B |  |  |  |  |  | p-values |  |  |
| --- | --- | --- | --- | --- | --- | --- | --- | --- | --- | --- | --- | --- | --- | --- | --- |
|  | Prescribed |  | Incidental |  | None |  | Prescribed |  | Incidental |  | None |  | Group*site | Group | Site |
|  | N | Value | N | Value | N | Value | N | Value | N | Value | N | Value |  |  |  |
| Age (years) | 121 | 71.4 (13.0) | 99 | 72.3 (13.9) | 31 | 73.9 (14.5) | 33 | 59.8 (14.4) | 12 | 65.8 (14.3) | 208 | 68.9 (13.9) | 0.21 | 0.0096 | <0.0001 |
| Sex |  |  |  |  |  |  |  |  |  |  |  |  |  |  |  |
| Female | 121 | 58 (47.9) | 99 | 48 (48.5) | 31 | 17 (54.8) | 33 | 10 (30.3) | 12 | 5 (41.7) | 208 | 83 (39.9) | - | 0.58 | 0.020 |
| Male |  | 63 (52.1) |  | 51 (51.5) |  | 14 (45.2) |  | 23 (67.7) |  | 7 (58.3) |  | 125 (60.1) |  |  |  |
| Time post-stroke (days) | 121 | 16.1 (16.2) | 99 | 18.2 (17.3) | 31 | 21.1 (15.1) | 33 | 22.4 (26.8) | 12 | 48.5 (46.9)* | 208 | 21.3 (23.3) | 0.0009 | - | - |
| Type of stroke |  |  |  |  |  |  |  |  |  |  |  |  |  |  |  |
| Ischemic | 121 | 91 (75.2) | 99 | 67 (67.7) | 31 | 25 (80.7) | 33 | 25 (75.8) | 12 | 8 (66.7) | 208 | 148 (71.2) | - | 0.46 | 0.029 |
| Lacune |  | 1 (0.8) |  | 1 (1.0) |  | 0 (0) |  | 0 (0) |  | 0 (0) |  | 0 (0) |  |  |  |
| Hemorrhagic |  | 19 (15.7) |  | 22 (22.2) |  | 2 (6.5) |  | 7 (21.2) |  | 2 (16.7) |  | 39 (18.8) |  |  |  |
| Transient ischemic attack |  | 1 (0.8) |  | 0 (0) |  | 0 (0) |  | 0 (0) |  | 0 (0) |  | 0 (0) |  |  |  |
| Unknown |  | 3 (2.5) |  | 2 (2.0) |  | 2 (6.5) |  | 0 (0) |  | 0 (0) |  | 0 (0) |  |  |  |
| Other |  | 6 (5.0) |  | 7 (7.1) |  | 2 (6.5) |  | 1 (3.0) |  | 2 (16.7) |  | 21 (10.1) |  |  |  |
| Affected hemisphere |  |  |  |  |  |  |  |  |  |  |  |  | - |  |  |
| Left | 121 | 54 (44.6) | 99 | 48 (48.5) | 31 | 12 (38.7) | 33 | 14 (42.4) | 12 | 8 (66.7) | 208 | 94 (45.2) |  | 0.83 | 0.69 |
| Right |  | 49 (40.5) |  | 39 (39.4) |  | 15 (48.4) |  | 17 (51.5) |  | 2 (16.7) |  | 84 (40.4) |  |  |  |
| Both |  | 15 (12.4) |  | 10 (10.1) |  | 4 (12.9) |  | 2 (6.1) |  | 2 (16.7) |  | 28 (13.5) |  |  |  |
| Unknown |  | 3 (2.5) |  | 2 (2.0) |  | 0 (0) |  | 0 (0) |  | 0 (0) |  | 2 (1.0) |  |  |  |
| More affected side |  |  |  |  |  |  |  |  |  |  |  |  | - |  |  |
| Left | 121 | 45 (37.2) | 99 | 43 (43.4) | 31 | 15 (48.4) | 33 | 16 (48.5) | 12 | 4 (33.3) | 208 | 92 (44.2) |  | 0.28 | 0.072 |
| Right |  | 49 (40.5) |  | 46 (46.5) |  | 12 (38.7) |  | 14 (42.4) |  | 7 (58.3) |  | 81 (38.9) |  |  |  |
| Both |  | 21 (17.4) |  | 10 (10.1) |  | 4 (12.9) |  | 1 (3.0) |  | 0 (0) |  | 23 (11.1) |  |  |  |
| Neither |  | 6 (5.0) |  | 0 (0) |  | 0 (0) |  | 3 (6.1) |  | 1 (8.3) |  | 12 (5.8) |  |  |  |
| Length of stay (days) | 121 | 33.3 (21.1) | 99 | 35.7 (22.2) | 31 | 39.6 (24.8) | 33 | 39.8 (15.9) | 12 | 39.5 (22.7) | 208 | 34.7 (16.2) | 0.091 | 0.95 | 0.50 |
| FIM total score | 121 | 66.0 (16.9) | 99 | 65.8 (14.4) | 31 | 58.8 (18.2)<sup>s</sup> | 33 | 67.8 (15.7) | 12 | 55.3 (25.8) | 208 | 68.4 (20.1)<sup>a</sup> | 0.0076 | - | - |
| FIM motor score | 120 | 41.1 (13.7) | 97 | 41.1 (12.3) | 30 | 33.5 (16.2)<sup>a</sup> | 33 | 43.3 (14.8) | 12 | 35.8 (22.1) | 208 | 45.8 (17.9)<sup>a</sup> | 0.0041 | - | - |
| FIM cognitive score | 120 | 25.1 (6.2) | 97 | 24.4 (6.0) | 30 | 25.7 (6.4) | 33 | 24.5 (6.4) | 12 | 19.5 (7.5) | 208 | 22.6 (5.7) | 0.11 | 0.034 | 0.0007 |
| FAC score | 121 | 2.5 (1.4)<sup>a</sup> | 99 | 2.0 (1.3) | 31 | 1.5 (1.8)<sup>a</sup> | 33 | 1.8 (1.4) | 12 | 1.5 (1.9) | 208 | 2.1 (1.5) | 0.0033 | - | - |
| CMSA arm score | 99 | 4.1 (1.2) | 84 | 4.0 (1.3) | 27 | 3.7 (2.0) | 27 | 3.3 (1.4) | 10 | 2.9 (1.8) | 141 | 3.9 (1.6) | 0.033 | - | - |
| CMSA hand score | 98 | 4.3 (1.3) | 85 | 4.1 (1.3) | 27 | 4 (1.9) | 27 | 3.5 (1.6) | 10 | 3.1 (1.6) | 140 | 4.1 (1.6) | 0.085 | 0.33 | 0.0083 |
| CMSA leg score | 101 | 4.4 (1.4) | 85 | 4.1 (1.3) | 24 | 3.6 (1.7) | 21 | 3.9 (1.5) | 8 | 3 (0.9) | 104 | 4 (1.2) | 0.019 | - | - |
| CMSA foot score | 99 | 4 (1.2) | 85 | 3.9 (1.4) | 22 | 3.2 (1.4) | 22 | 3.5 (1.6) | 8 | 3.5 (1.4) | 103 | 3.7 (1.4) | 0.098 | 0.45 | 0.56 |

\*The Incidental group at Site B was significantly higher than all other group & site combinations.

Data for primary outcomes at discharge from rehabilitation are shown in Figure 1. There was a significant site-by-group interaction effect for FIM total score (F_2,495_=8.49; p=0.0002). Post-hoc testing revealed that, at Site A, the Prescribed and Incidental groups had higher FIM total scores at discharge than the None group, and that the None group at Site B had higher FIM total scores at discharge than the None group at Site A. Regarding secondary outcomes, there were significant group-by-site interaction effects for FIM motor sub-score (F_2,495_=12.84; p<0.0001), FAC (F_2,495_=8.24; p=0.0003), and CMSA hand scores (F_2,495_=3.38; p=0.036). Post-hoc analysis showed that the Prescribed group had higher FIM and FAC scores at discharge than either the Incidental group (Site A for FIM motor scores) or the None group (Site B for both FIM motor scores and FAC scores). While there was a significant group-by-site interaction effect for CMSA hand scores, there were no statistically significant comparisons at p<0.05 on post-hoc testing. There was no significant group-by-site interaction effect for FIM cognitive sub-score (F_2,495_=0.35; p=0.71) or CMSA leg scores (F_2,495_=1.32; p=0.27), but there were significant main effects for group (FIM cognitive: F_2,495_=3.13; p=0.045; CMSA leg: F_2,495_=4.52; p=0.012); post-hoc testing revealed that the Prescribed group had significantly higher FIM cognitive scores and CMSA leg scores at discharge than the None group, across both sites combined. There were no significant group-by-site interaction effects (F_2,495_<2.71; p>0.068) or main effects of group (F_2,495_<1.39; p>0.25) for CMSA arm or foot scores.

**Figure 1:**
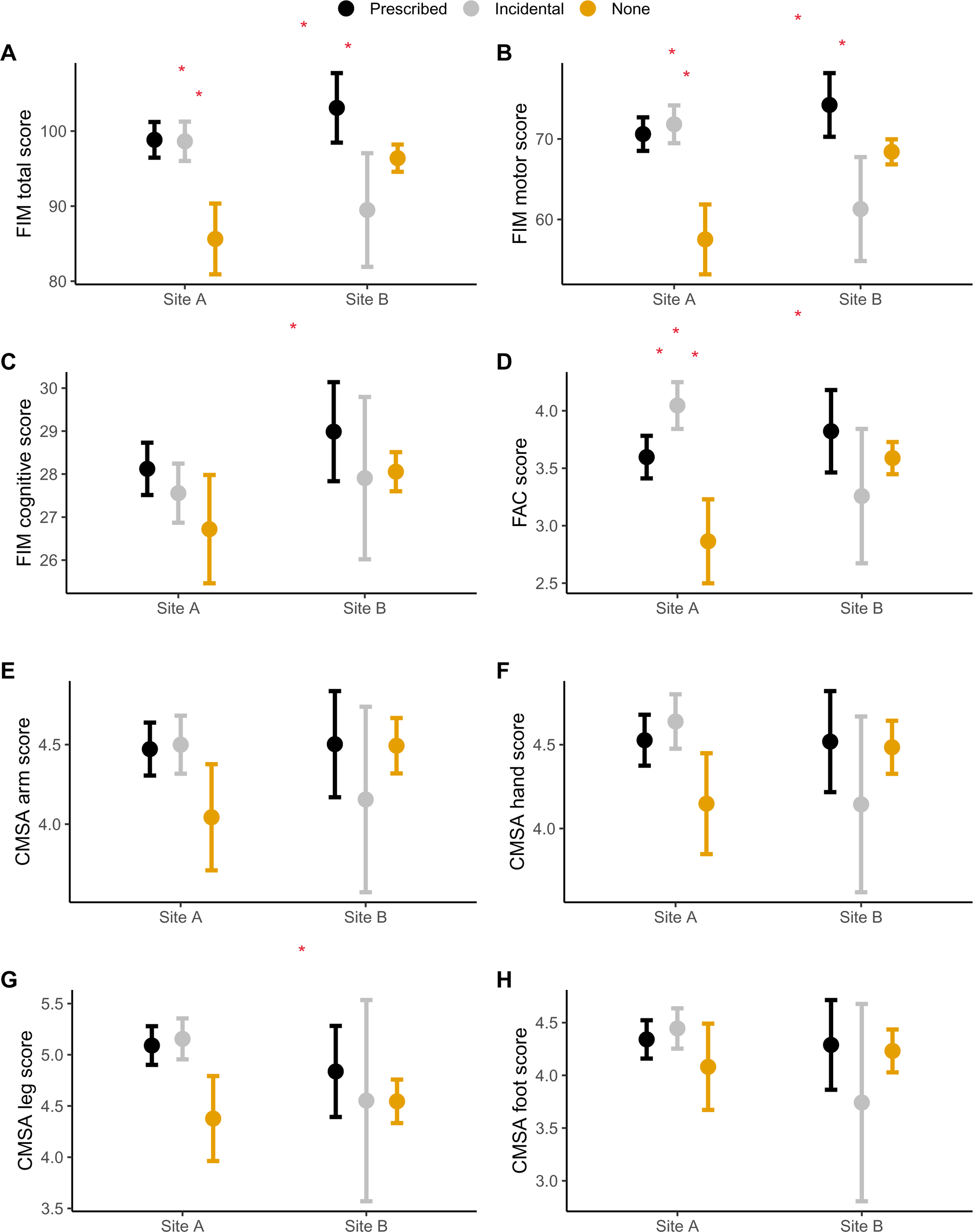
Outcomes at discharge from rehabilitation by group and site. Values plotted are least square means controlling for other effects in the model (outcome values at admission, length of stay, and age) with standard deviation error bars for FIM total score (Panel A), FIM motor sub-score (Panel B), FIM cognitive sub-score (Panel C), FAC score (Panel D), CMSA arm score (Panel E), CMSA hand score (Panel F), CMSA leg score (Panel G), and CMSA foot score (Panel H). Values joined by square brackets and marked with an asterisk are statistically significantly different from each other (p<0.05).

Figure 2 displays results of regression analysis of time spent doing CRE and change in outcomes. There were statistically significant positive correlations for all outcomes, except for CMSA foot and hand scores. That is, more time spent exercising was correlated with greater improvement in FIM total and motor and cognitive sub-scores, FAC and CMSA arm and leg scores. For illustration, regression revealed 5 minutes of exercise per day corresponded to a 12-point increase in total FIM score over 30 days of inpatient rehabilitation.

**Figure 2:**
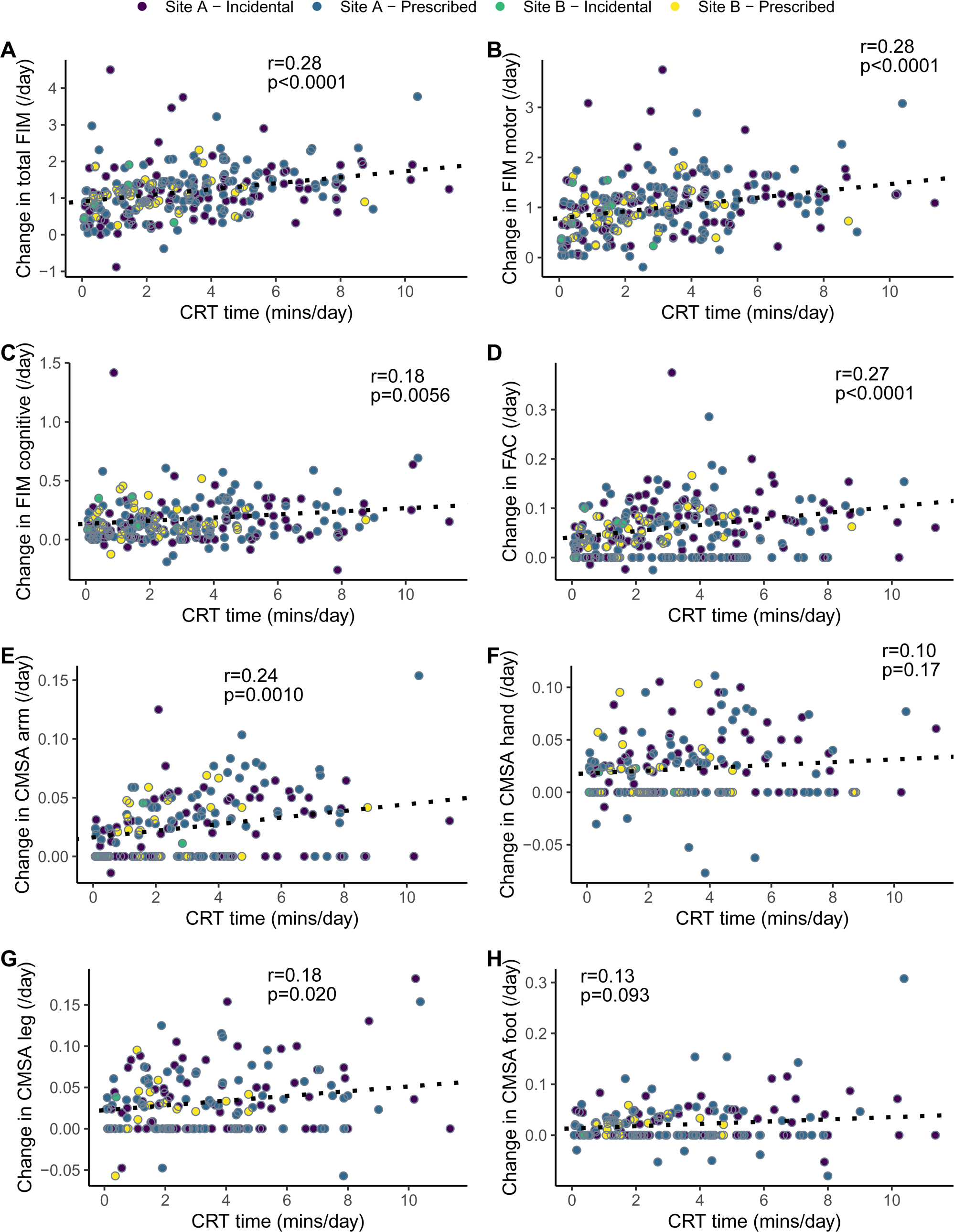
Scatter plots showing the relationship between time spent doing cardiorespiratory exercise (CRE) and change in outcomes. Values plotted are discharge minus admission score, divided by length of stay, for FIM total score (Panel A), FIM motor sub-score (Panel B), FIM cognitive sub-score (Panel C), FAC score (Panel D), CMSA arm score (Panel E), CMSA hand score (Panel F), CMSA leg score (Panel G), and CMSA foot score (Panel H). Pearson partial correlations (controlling for site and age), and associated p-values are shown for each comparison.

## DISCUSSION

The aim of this longitudinal observational cohort study was to determine if participation in CRE early post-stroke, during routine inpatient stroke rehabilitation, improves functional independence related to motor and cognitive activities (i.e., FIM scores), ability to ambulate independently (i.e., FAC scores), and recovery of stroke-related leg, foot, arm, and hand impairments (i.e., CMSA scores). Our primary hypothesis was that total FIM scores would be higher at discharge from in-patient stroke rehabilitation for those who completed CRE than those who did not complete CRE. There was some evidence to support this hypothesis; at Site A, patients who completed any CRE (either prescribed or incidentally implemented) had higher FIM scores than those who did not complete CRE, whereas at Site B FIM scores were only higher for those patients who were prescribed CRE compared to those who completed CRE without a prescription. Evidence from controlled trials indicates that CRE improves cerebral blood circulation, which in turn could enhance motor and cognitive function.^24^ Investigating the benefits of CRE through rehabilitation in early-stage stroke patients, as well as understanding its association with functional recovery in the post-acute phase of stroke, has proven challenging, with few clinical trials conducted to date.^3,8,25,26^ In the absence of large clinical trials, alternative study designs (e.g., observational studies like ours or quasi-experimental studies^27^) can provide evidence for the potential benefits of CRE for improving function early post-stroke. We found evidence that CRE early post-stroke (minimum 2 days post-stroke) during routine inpatient stroke rehabilitation stay (length of stay 8-114 days) can result in increased total FIM scores at discharge, particularly when CRE was included as part of the patients’ treatment plan (FIM scores on average 9.3 points higher for the Prescribed group than the None group at discharge, across both sites combined).

We also hypothesized that FIM motor and cognitive sub-scores would improve more for those who completed CRE during stroke rehabilitation than those who did not. Findings for the FIM motor sub-score were similar to the FIM total score, which may reflect the fact that the FIM motor sub-score is a larger component of the total score than the cognitive sub-score, or that CRE, which involves motor exercise, primarily improves motor function. However, it is noteworthy that FIM cognitive sub-scores were higher at discharge from in-patient stroke rehabilitation for the Prescribed group than the None group. Others have found that participating in CRE early post-stroke improved certain aspects of cognitive function, such as attention and processing speed,^28^ but not memory, problem solving, executive function, or working memory.^3,28^ In our study, communication and social cognition domains related to functional independence measured through FIM cognitive scores were noted to improve at the time of discharge from in-patient stroke rehabilitation among those who participated in CRE.

Our secondary hypotheses were that ambulation ability measured through FAC, and stroke-related motor impairment measured through CMSA leg, foot, arm, and hand scores would improve more for those who completed CRE during in-patient stroke rehabilitation than those who did not. For the FAC, higher scores at discharge were observed for both CRE groups compared to the None group at Site A only. Interestingly, the Incidental group had higher FAC scores at discharge than the Prescribed group. It is possible that physiotherapists at this site included CRE for patients who quickly recovered functional ambulation as they no longer needed to work on ambulatory activities in therapy sessions.^10^ Similar to FIM motor sub-scores, increased CMSA leg scores (but not arm, hand, or foot scores), for those who were prescribed CRE compared to those who did no CRE may reflect specificity of training. That is, because CRE involves repeated movement, typically of the large muscle groups in the lower extremity, benefits are more likely for lower-extremity gross motor control than upper extremity or fine (i.e., foot) motor control.^14^

Providing further evidence for a possible link between CRE and improved outcomes, we found that higher total time spent in cardiorespiratory exercise correlated with greater improvement in total FIM, FIM motor and cognitive sub-scores, FAC, and CMSA arm and leg scores. In agreement with our results, Nozoe et al.^29^ reported moderate positive correlations between CRE time and FIM motor scores at discharge in the patients with stroke who were ambulatory, and with FIM cognitive score in those who were non-ambulatory. These results along with our findings indicate that a higher dose (i.e., increased duration) of CRE could lead to greater gains in functional independence.

### Limitations

This is a longitudinal observational cohort study, and therefore we can not infer causality from the findings, i.e., whether participation in CRE resulted in improved outcomes. It is possible that physiotherapists chose to complete CRE with their patients who were expected to make greater functional gains, whereas they had other rehabilitation priorities for those with expected slower gains in functional recovery.^10^ However, the groups did not differ on baseline FIM scores or length of stay. While the positive correlations between CRE time and outcomes provides some evidence for CRE possibly contributing to improved outcomes, it is also possible that physiotherapists progressed patients who recovered faster to complete a greater frequency and/or duration of CRE. CRE was completed as part of routine care, and we relied on physiotherapists’ documentation of CRE. Intensity of CRE was not often documented, particularly at Site A, so it was unclear if the intensity was optimal to improve cardiorespiratory fitness. Additionally, physiotherapists may not have comprehensively documented their treatment plans, or may have amended their treatment plan partway through patients’ length of stay, leading to some participants being misclassified in the Incidental group. Data were collected at two urban hospitals in Ontario, Canada. Some findings differed between the two sites, for example, whether the Prescribed group only, or both the Prescribed and Incidental groups combined, had improved outcomes at discharge. These differing findings may suggest differences in care practices between the two sites. The findings may not generalize to sites or regions with different care practices or models of care.

### Conclusion

This study investigated the effects of participation in CRE early post-stroke on recovery from stroke, via functional independence and safe ambulation. We observed a significantly higher FIM total scores, FIM motor and cognitive sub-scores, FAC scores, and CMSA leg scores for those who completed prescribed CRE compared to patients who did no CRE during routine stroke rehabilitation, when controlling for baseline scores and length of stay. Results of this study suggest that participation in CRE may improve functional independence for people with sub-acute stroke. These results will need to be confirmed with a more controlled study design.

## Data Availability

Raw data are not available externally due to privacy legislation.

## ACKNOWLEDGEMENTS

We thank Kay-Ann Allen and Alex Kalli for their assistance with data collection.

## REFERENCES

1. Ploughman M, Kelly LP. Four birds with one stone? Reparative, neuroplastic, cardiorespiratory, and metabolic benefits of aerobic exercise poststroke. Curr Opin Neurol. 2016;29(6):684–692.

2. Billinger SA, Arena R, Bernhardt J, et al. Physical activity and exercise recommendations for stroke survivors: a statement for healthcare professionals from the American Heart Association/American Stroke Association. Stroke. 2014;45:2532–2553.

3. Saunders DH, Sanderson M, Hayes S, et al. Physical fitness training for stroke patients. Cochrane Database Syst Rev. 2020;3(3):CD003316.

4. Rimmer JH, Wang E. Aerobic exercise training in stroke survivors. Top Stroke Rehabil. 2005;12(1):17–30.

5. Ploughman M, Austin MW, Glynn L, Corbett D. The effects of poststroke aerobic exercise on neuroplasticity: a systematic review of animal and clinical studies. Transl Stroke Res. 2015;6(1):13–28.

6. Austin MW, Ploughman M, Glynn L, Corbett D. Aerobic exercise effects on neuroprotection and brain repair following stroke: a systematic review and perspective. Neurosci Res. 2014;87:8–15.

7. Corbett D, Nguemeni C, Gomez-Smith M. How can you mend a broken brain? Neurorestorative approaches to stroke recovery. Cerebrovasc Dis. 2014;38:233–239.

8. Stoller O, de Bruin ED, Knols RH, Hunt JJ. Effects of cardiovascular exercise early after stroke: systematic review and meta-analysis. BMC Neurology. 2012;12:45.

9. Hebert D, Lindsay MP, McIntyre A, et al. Canadian stroke best practice recommendations: stroke rehabilitation practice guidelines, update 2015. Int J Stroke. 2016;11(4):459–484.

10. Barzideh A, Devasahayam AJ, Tang A, et al. Physiotherapists’ use of aerobic exercise during stroke rehabilitation: a qualitative study using chart-stimulated recall. medRxiv. 2023;doi:10.1101/2023.12.13.23299927.

11. Kendall BJ, Gothe NP. Effect of aerobic exercise interventions on mobility among stroke patients: a systematic review. Am J Phys Med Rehabil. 2016;95(3):214–224.

12. Zheng G, Zhou W, Xia R, Tao J, Chen L. Aerobic exercises for cognition rehabilitation following stroke: a systematic review. J Stroke Cerebrovasc Dis. 2016;25(11):2780–2789.

13. Bernhardt J, Hayward KS, Kwakkel G, et al. Agreed definitions and shared vision for new standards in stroke recovery research: the stroke recovery and rehabilitation roundtable taskforce. Int J Stroke. 2017;12(5):444–450.

14. Kwakkel G, Wagenaar RC, Twisk JWR, Lankhorst GJ, Koetsier JC. Intensity of leg and arm training after primary middle-cerebral-artery stroke: a randomised trial. Lancet. 1999;354:191–196.

15. Chumney D, Nollinger K, Shesko K, Skop K, Spencer M, Newton RA. Ability of Functional Independence Measure to accurately predict functional outcome of stroke-specific population: systematic review. J Rehabil Res Dev. 2010;47(1):17–29.

16. Holden MK, Gill KM, Magliozzi MR, Nathan J, Piehl-Baker L. Clinical gait assessment in the neurologically impaired: reliability and meaningfulness. Phys Ther. 1984;64(1):35–40.

17. Gowland C, Stratford P, Ward M, et al. Measuring physical impairment and disability with the Chedoke-McMaster Stroke Assessment. Stroke. 1993;24:58–63.

18. Canadian Institute for Health Information. Rehabilitation. 2019; https://www.cihi.ca/en/rehabilitation. Accessed 31 July, 2019.

19. Health Quality Ontario, Care MoHaL-T. Quality-based procedures: clinical handbook for stroke (acute and postacute). Toronto 2016.

20. Evensen J, Soberg HL, Sveen U, Hestad KA, Moore JL, Bronken BA. Individualized goals expressed by patients undergoing stroke rehabilitation: an observational study. J Rehabil Med. 2024;56:jrm15305.

21. Hasan SMM, Rancourt SN, Austin MW, Ploughman M. Defining optimal aerobic exercise parameters to affect complex motor and cognitive outcomes after stroke: a systematic review and synthesis. Neural Plasticity. 2016;2016:2961573.

22. Vandenbroucke JP, von Elm E, Altman DG, et al. Strengthening the reporting of observational studies in epidemiology (STROBE): explanation and elaboration. PLoS Med. 2007;4(10):e297.

23. Canadian Institutes of Health Research, Natural Sciences and Engineering Research Council of Canada, Social Sciences and Humanities Research Council of Canada. Tri-Council Policy Statement: Ethical Conduct for Research Involving Humans. December 2022.

24. Padilla J, Simmons GH, Bender SB, Arce-Esquivel AA, Whyte JJ, Laughlin MH. Vascular effects of exercise: endothelial adaptations beyond active muscle beds. Physiology. 2011;26(3):132–145.

25. Katz-Leurer M, Carmeli E, Shochina M. The effect of early aerobic training on independence six months post stroke. Clin Rehabil. 2003;17:735–741.

26. Gezer H, Karaahmet OZ, Gurcay E, Dulgeroglu D, Cakci A. The effect of aerobic exercise on stroke rehabilitation. Ir J Med Sci. 2019;188(2):469–473.

27. Henderson CE, Plawecki A, Lucas E, et al. Increasing the amount and intensity of stepping training during inpatient stroke rehabilitation improves locomotor and non-locomotor outcomes. Neurorehabil Neural Repair. 2022;36(9):621–632.

28. Oberlin LE, Waiwood AM, Cumming TB, Marsland AL, Bernhardt J, Erickson KI. Effects of physical activity on poststroke cognitive function: a meta-analysis of randomized controlled trials. Stroke. 2017;48(11):3093–3100.

29. Nozoe M, Masuya R, Yamamoto M, Kubo H, Kanai M, Shimada S. Correlations between aerobic exercise time during physiotherapy and characteristics of patients with subacute stroke: a pilot cros-sectional study. Physiother Theory Pract. 2023;39(2):433–440.

